# Early Detection of Novel SARS-CoV-2 Variants from Urban and Rural Wastewater through Genome Sequencing and Machine Learning

**DOI:** 10.1101/2024.04.18.24306052

**Authors:** Xiaowei Zhuang, Van Vo, Michael A. Moshi, Ketan Dhede, Nabih Ghani, Shahraiz Akbar, Ching-Lan Chang, Angelia K. Young, Erin Buttery, William Bendik, Hong Zhang, Salman Afzal, Duane Moser, Dietmar Cordes, Cassius Lockett, Daniel Gerrity, Horng-Yuan Kan, Edwin C. Oh

## Abstract

Genome sequencing from wastewater has emerged as an accurate and cost-effective tool for identifying SARS-CoV-2 variants. However, existing methods for analyzing wastewater sequencing data are not designed to detect novel variants that have not been characterized in humans. Here, we present an unsupervised learning approach that clusters co-varying and time-evolving mutation patterns leading to the identification of SARS-CoV-2 variants. To build our model, we sequenced 3,659 wastewater samples collected over a span of more than two years from urban and rural locations in Southern Nevada. We then developed a multivariate independent component analysis (ICA)-based pipeline to transform mutation frequencies into independent sources with co-varying and time-evolving patterns and compared variant predictions to >5,000 SARS-CoV-2 clinical genomes isolated from Nevadans. Using the source patterns as data-driven reference “barcodes”, we demonstrated the model’s accuracy by successfully detecting the Delta variant in late 2021, Omicron variants in 2022, and emerging recombinant XBB variants in 2023. Our approach revealed the spatial and temporal dynamics of variants in both urban and rural regions; achieved earlier detection of most variants compared to other computational tools; and uncovered unique co-varying mutation patterns not associated with any known variant. The multivariate nature of our pipeline boosts statistical power and can support accurate and early detection of SARS-CoV-2 variants. This feature offers a unique opportunity for novel variant and pathogen detection, even in the absence of clinical testing.

## Introduction

Public health testing and the sequencing of SARS-CoV-2 genomes have been pivotal in advancing the development of precise vaccine therapeutics and facilitating comprehensive surveillance of circulating variants^1,2^. However, clinical surveillance of COVID-19 transmission often imposes substantial demands on laboratory resources and relies on individuals actively seeking testing^3,4^. These challenges underscore the need for complementary, proactive, and cost-effective methods to monitor the emergence and spread of novel SARS-CoV-2 variants.

In the United States, there has been substantial spatial and temporal variation reported for the dynamics of the COVID-19 pandemic in urban and rural counties^5,6^. Studies have described disease incidence being initially high in urban locations, followed by a rapid surge in infections from rural areas^5,6^. Notably, the overall disease incidence is reported to be lower in rural compared to urban regions^5,7^. Given that rural communities have fewer healthcare resources and an established reluctance to seek medical care, an overarching infection prevention and control effort could benefit significantly from new interventions designed to track disease incidence.

Wastewater-based epidemiology (WBE) has emerged as a valuable alternative for tracking changes in SARS-CoV-2 viral levels and variants within a community^8–12^. The SARS-CoV-2 virus is a single-stranded RNA virus that can be shed in wastewater through human waste such as feces, saliva, and urine^13,14^. In comparison to clinical testing, wastewater analyses provide a less-biased approach to viral monitoring, particularly in areas with limited healthcare resources and unclear testing hesitancy rates^15–19^. Throughout the COVID-19 pandemic, WBE served as a pivotal and cost-effective tool to monitor and characterize the emergence and spread of SARS-CoV-2 variants of concerns (VoCs), offering early detection for potential outbreaks^8,18,20–23^.

The complexity of wastewater matrices presents significant challenges in obtaining high-quality nucleic acid sequences and detecting SARS-CoV-2 variants. To overcome this shortfall, targeted hybridization and amplicon-based sequencing methods have been implemented to characterize the viral composition within a sample^24–31^. Complementing these sequencing approaches, bioinformatic pipelines have then been developed with unique computational considerations that support the identification and quantification of SARS-CoV-2 variants^18,21,32–36^. For example, the COJAC pipeline^33^ utilizes a variant-specific mutation pattern and counts aligned read pairs to detect the presence of a VoC in wastewater samples, even with a relatively low viral load. In addition, the Vpipe^37^, LoFreq^38^, or iVar^39^ pipeline can be used to define single nucleotide polymorphisms (SNPs) with alternative allele frequencies within the SARS-CoV-2 genome. To determine the presence and abundance of VoCs, these pooled SNP alternative allele frequencies are generally modeled as a linear combination of predefined VoCs using GISAID or UshER-curated reference barcodes^20,21,34–36^. Among various linear regression models and optimization methods to estimate the abundance of variants, a pipeline called Freyja^34^ is frequently employed due to its simplicity in modeling and interpretation. However, despite the widespread use of these pipelines, a shared bias toward pre-defined reference barcodes exists, potentially leading to incorrect variant predictions with metadata errors. This bias can be exacerbated when either 1) the variants included in the reference barcodes do not match the current circulating VoCs in the communities, or 2) a new VoC may be circulating within the community, but has not been identified through clinical sequencing. Furthermore, since wastewater samples represent a composite of multiple clinical genomes, there may be limited statistical power to detect emerging VoCs with low abundance in a single sample.

Here, we introduce a multivariate method designed to analyze SARS-CoV-2 wastewater sequencing data and identify circulating variants. We hypothesize that our independent component analysis (ICA)-based pipeline called ICA-Var (Independent Component Analysis of Variants) can leverage multiple sequencing datasets to amplify statistical power and thereby enable early and precise detection of variants within the community. To validate our approach, we compare the results obtained using our pipeline with those generated by the state-of-the-art tool Freyja^34^. Our approach also identifies emerging co-varying mutation patterns, which may belong to more recent VoCs or have not been reported. Collectively, our findings demonstrate the effectiveness of this new pipeline, even in the absence of clinical data. These results underscore the potential for ICA-Var to identify mutation patterns within the SARS-CoV-2 genome that could give rise to novel circulating variants.

## Results

### Large-scale genome sequencing and the development of a computational pipeline

Analyzing wastewater SARS-CoV-2 genomes presents inherent complexities resulting from various factors. For example, the use of short sequencing reads introduces challenges in accurately phasing genomes and the degradation of viral genomes in environmental samples contributes to uneven genome coverage and sequencing read depth. To address these challenges, we sequenced 3,659 wastewater samples using an amplicon-targeted approach and employed stringent quality control measures. Using a minimum threshold of 80% genome coverage at more than 50X sequencing depth, we selected 1,385 of these samples, covering 59,422 locations/mutations on the genome, for further analysis (**Supplementary Figure 1**). Leveraging this extensive dataset, we developed a data-driven approach named ICA-Var. This method transforms mutation frequencies in wastewater samples into independent sources with co-varying mutation patterns and utilizes a dual-regression method to re-associate the independent sources back to the original samples (**Figure 1A and Methods**). We hypothesized that the ICA sources could effectively capture the evolving dominant SARS-CoV-2 variants over time, with each being characterized by distinct determinant mutation patterns. To evaluate the performance of our tool, we conducted a comparative analysis of variant detection against the state-of-the-art tool known as Freyja (**Figure 1A-B**).

**Figure 1.**
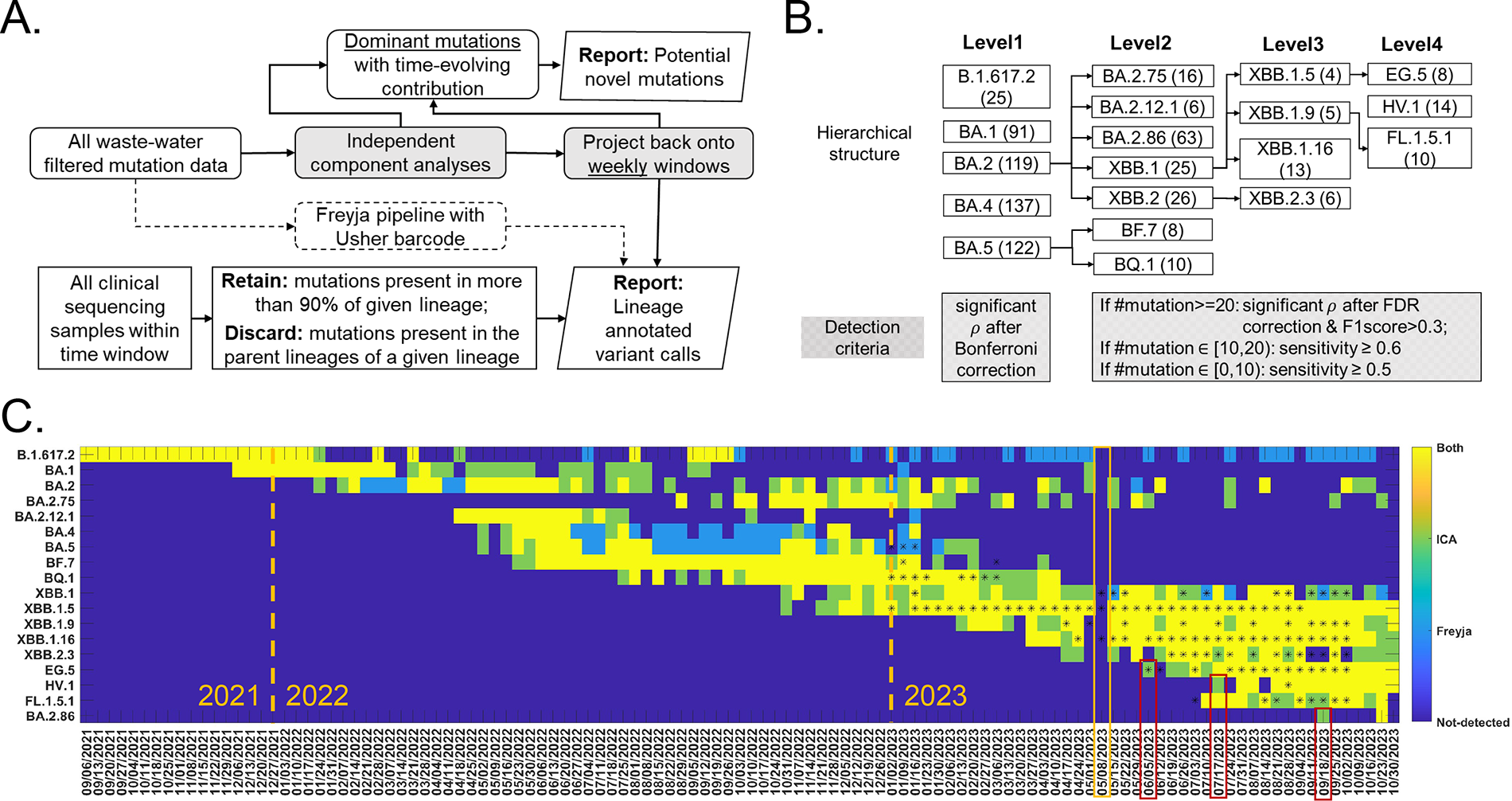
ICA-Var pipeline and comparisons with Freyja. **(A)** Proposed independent component analyses (ICA) pipeline. Two matrices are reported: SARS-CoV-2 lineages detection each week (bottom row), and potential novel mutations (top row). **(B)** Hierarchical structure of 18 variants of concerns (VoCs). Lineage-defining mutations for each VoC were obtained from clinical data summarized at covspectrum.org, and the number of defining mutations were listed in brackets. Criteria for calling a detection in the proposed pipeline were listed in shaded boxes. Abbreviations: ρ: the Spearman’s Correlation coefficient; FDR: false discovery rate. **(C).** Detection of the emerging VoCs in wastewater from Southern Nevada from August 2021 to November 2023 in the proposed method (first reporting matrix in **(A)**) and the state-of-art tool Freyja. An asterisk (*) indicates at least one clinical sample was reported within that week. Earlier detections of the proposed method were observed for emerging variants EG. 5, HV.1, and BA.2.86 (red triangle boxes). The yellow triangle box indicates the week without wastewater sampling due to technical issues.

In late 2021, both Freyja and ICA-Var reliably identified B.1.617.2 (Delta) and BA.1 (Omicron) VoCs in wastewater samples (**Figure 1C**, yellow in first two rows), reflecting the prevalence of both variants during this period. In 2022, ICA-Var demonstrated the ability to detect BA.2, BA.4, BA.5, BF.7, BQ.1, XBB.1, and XBB.1.5 variants one or several weeks before Freyja (**Figure 1C**, green). Consistent detection of Omicron variants was obtained by Freyja and ICA-Var throughout 2022 (**Figure 1C**, yellow). In 2023, both Freyja and ICA-Var successfully identified XBB.1.16 in late March, a month prior to the first sequenced clinical sample in Southern Nevada (**Figure 1C**, yellow) For more emerging VoCs in 2023, such as EG.5, ICA-Var detected this variant in early June, coinciding with the week of the first reported clinical sequence in Southern Nevada (first red box in **Figure 1C**). In contrast, Freyja reliably identified the EG.5 signal only once the VoC became more prevalent in early July. Similarly, for HV.1, and BA.2.86 VoCs, ICA-Var detected the presence of these variants in wastewater several weeks before Freyja (**Figure 1C**, red boxes).

To explore the earlier detection of the emerging VoCs EG.5, HV.1, and BA.2.86 by ICA-Var compared to Freyja, we generated a heatmap illustrating alternative allele frequencies at the dominant mutation sites for these variants. This heatmap represents samples from the first week of detection by each method (**Figure 2**). Specifically, for EG.5, ICA-Var initially identified the variant during the week of 06/05/2023, and two wastewater samples in this week exhibited reliable mutation frequencies at three out of eight EG.5 dominant mutation sites (**Figure 2**, top row in panel EG.5). Freyja reported abundances of 0.66% and 0.53% for EG.5 in these two samples, respectively. Furthermore, an additional wastewater sample from the same week showed reliable mutation frequencies at two EG.5 dominant mutation sites, but Freyja did not identify EG.5 in this particular sample. As a multivariate method, ICA-Var leveraged all these samples with reliable, yet relatively low prevalence of EG.5 mutation sites, thereby enhancing statistical power and consequently enabling an earlier detection of EG.5 in this particular week. Conversely, Freyja reported a 23.08% abundance of one wastewater sample in the week of 07/10/2023. As shown in **Figure 2** (bottom row in panel EG.5), this sample demonstrated reliable alternative allele frequencies at five out of eight EG.5 determinant mutation sites (**Figure 2**, red box). The increased prevalence of EG.5 mutation sites in this sample contributed to the detection of EG.5 in Freyja. Similarly, for HV.1 (**Figure 2**, HV.1 panel) and BA.2.86 (**Figure 2**, BA.2.86 panel), ICA-Var capitalized on multiple wastewater samples with reliable yet relatively low prevalence of dominant mutation sites, thereby enhancing statistical power and achieving an earlier detection. In contrast, Freyja mandated at least one individual sample to exhibit the presence of dominant mutation sites for detection (red boxes in **Figure 2**).

**Figure 2.**
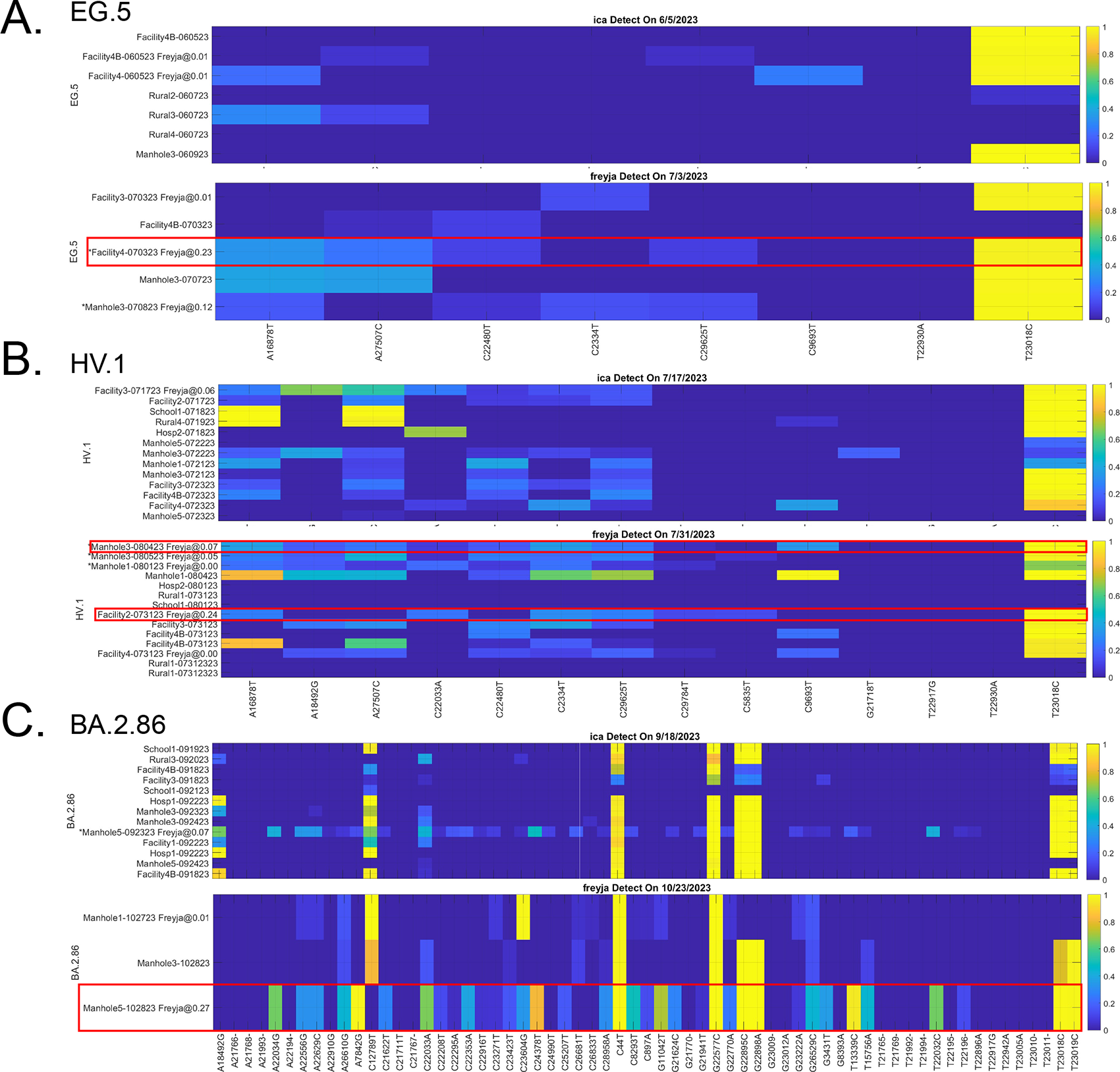
Detection of three variants using ICA-Var and Freyja. **(A)** Earlier detection of EG.5, **(B)** HV.1, and **(C)** BA.2.86 made by the proposed method, as compared to Freyja. In each panel, top and bottom rows plot the alternative allele frequencies at the dominant mutation sites for samples at the first detection date made by the proposed method and Freyja, respectively. X-axis represents each determinant mutation for the variant, y-axis represents each individual sample at the first detection date, and the color represents the alternative allele frequencies. In the y-axis, for each sample, if the Freyja pipeline outputs an abundance, the abundance value will be listed after the sample name. An asterisk (*) before the sample name indicates that the variant could be detected by this sample following the proposed criteria.

We further evaluated the earlier detection of VoCs in 2022 for ICA-Var and Freyja (**Supplementary Figure 2**). In addition to enhancing statistical power, the inclusion of deletions as additional sites in the proposed pipeline (indicated by orange boxes in **Supplementary Figure 2**) played a significant role in the earlier detections of BA.2, BA.4, and BA.5 variants compared to Freyja. This advantage arises from the fact that no deletions were utilized in the inference process within the default settings of Freyja, a result previously discussed in another computational pipeline designed to analyze wastewater sequencing data^35^ .

### Detection of VoCs in urban and rural samples

From the beginning of 2022, we sequenced and analyzed wastewater samples from rural areas in Southern Nevada (**Supplementary Table 1**). The number of urban (orange curve) and rural (yellow curve) samples for each week were plotted in **Supplementary Figure 1D**. We conducted a comprehensive urban-rural epidemiological comparison in our wastewater analyses (**Methods**), with samples categorized as urban and rural analyzed separately for each week. We present a summary of the detection of 18 VoCs utilizing both the established Freyja pipeline (**Figure 3A**) and our proposed ICA-Var pipeline. (**Figure 3B**).

**Figure 3.**
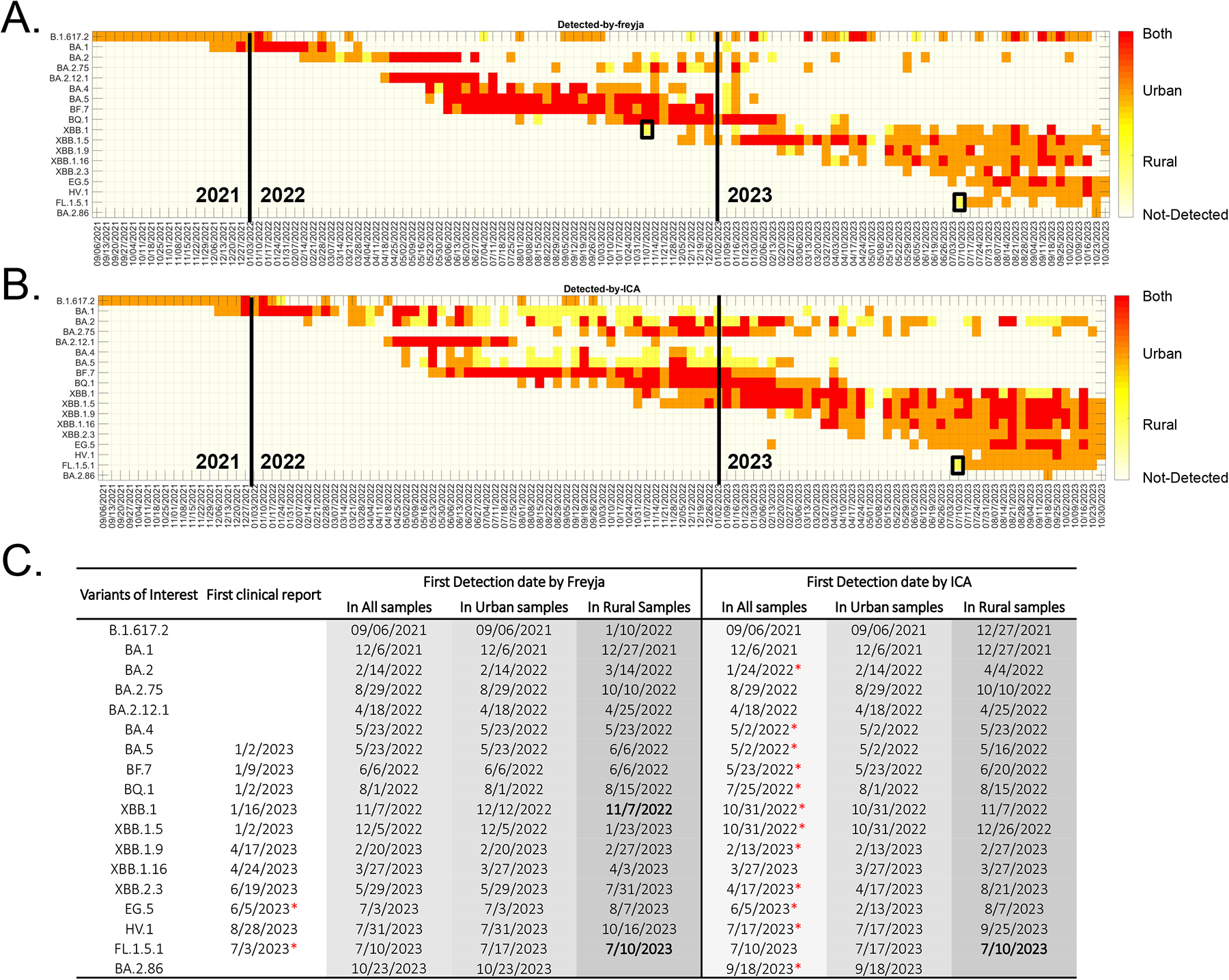
Variant detection in urban and rural samples. **(A-B)** Detection of the emerging variants of concerns (VoCs) in urban and rural samples using **(A)** Freyja and the **(B)** ICA-Var pipeline. **(C)** Earliest date of variant detection in clinical cases from Southern Nevada; first detection date with all wastewater samples (lightest grey), urban wastewater samples (lighter grey) and rural wastewater samples (darker grey) using Freyja and the proposed pipeline. A red asterisk (*) indicates the earliest detection dates among clinical reports, Freyja, and the proposed method. If the variant was captured by the Freyja and ICA-Var on the same earliest date, no * would be indicated. Earlier detection dates in rural samples than urban samples are highlighted in bold.

ICA-Var and Freyja both identified 16 out of the 18 VoCs in urban wastewater samples prior to detecting these VoCs in wastewater samples from rural locations (**Figure 3A-B**). This data suggest that new SARS-CoV-2 variants typically enter urban areas first before spreading into rural areas. Interestingly, XBB.1 and FL.1.5.1 were first detected in rural wastewater samples by either Freyja or ICA-Var (black boxes in **Figure 3A-B**). More specifically, Freyja first identified the XBB.1 variant in a rural sample in the week of 11/07/2022 (dashed red box in **Supplementary Figure 2**, panel XBB.1), but ICA-Var was able to detect XBB.1 one week prior to Freyja in urban samples (**Figure 3C**). Both ICA-Var and Freyja first detected FL.1.5.1 in rural samples on 07/10/2023 (**Figure 3C**). Detailed inspections showed that one rural sample on 07/12/2023 showed an overwhelming presence of FL.1.5.1 dominant mutations (dashed red box in **Supplementary Figure 2**, panel FL.1.5.1), which contributed to this earlier detection in rural areas. In contrast, urban samples demonstrated a much lower alternative allele frequencies and prevalence at FL.1.5.1 mutations.

### Identification of mutation sites with significant time-evolving contributions

Out of 59,422 mutation sites included, and following the analyses pipeline in **Figure 1A**, a total of 730 mutation sites demonstrated significant contributions during the multivariate group ICA (**Supplementary Figure 1B**). Among them, a subset of 177 mutations showed a significant time-evolving contribution from August 2021 to November 2023 **(Methods)**. As a proof of concept, we cross-referenced these 177 mutations with dominant mutation sites in B.1.617.2, BA. 1 and XBB.1 variants, and plotted their weekly contributions in **Figure 4**.

**Figure 4.**
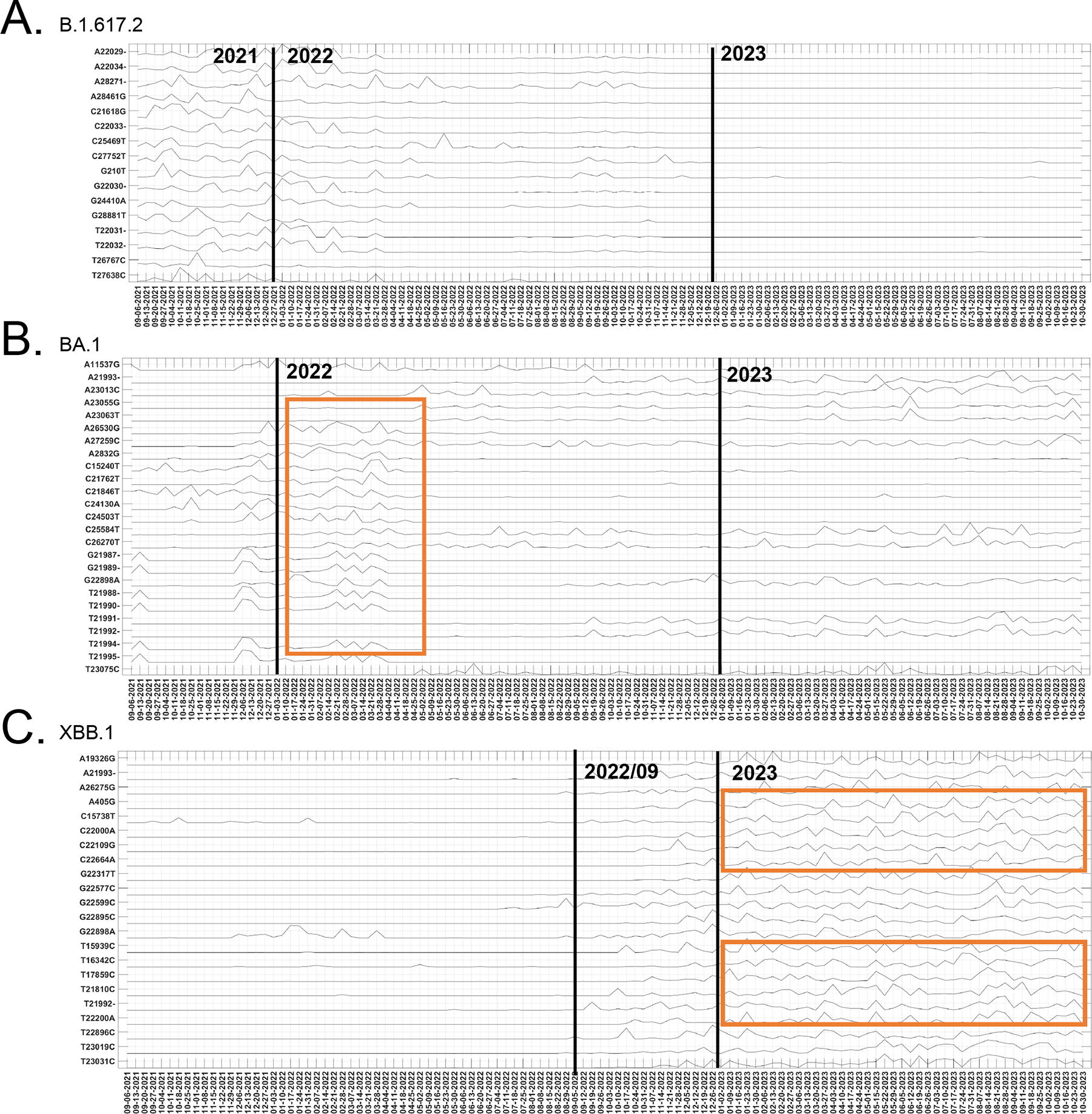
Mutations with significant time-evolving contributions in the proposed method. **(A)** Following the proposed method, 16 out of 25 determinant mutations in B.1.617.2 were identified to maintain a significant time-evolving contributions to the group, with major contributions in 2021 and early 2022. **(B)** The 25 dominant mutations in BA.1 were identified to have significant time-evolving contributiong to the group, with major contributions in early 2022. **(C)** We identified 22 out of 25 determinant mutations in XBB.1 to have significant time-evolving contributions to the group, with major contributions after 2022/09. Similar contributing patterns were observed for several determinant mutations (orange boxes).

Significant fluctuating contributions were observed in late 2021 for 16 out of 25 dominant mutation sites in B.1.617.2 (**Figure 4**, panel B.1.617.2). These contributions gradually declined through 2022 and diminished further in 2023. For the BA.1 variant, there was a noticeable increase in contributions related to the associated mutations in late 2021, peaking in early 2022 (**Figure 4**, panel BA.1, orange box). For several BA.1 mutation sites, time-evolving contributions continued to fluctuate in 2023, and their involvement in other Omicron sub-lineages (e.g., XBB.1) was reported at nextstrain.org. In addition, 22 out of 25 dominant mutations in XBB.1 displayed significant time-evolving contributions, with a substantial impact after September 2022. Similar fluctuation patterns were observed for several mutation sites (**Figure 4**, panel XBB.1, orange box), indicating that these mutation sites co-vary together and demonstrate a recombinant nature for XBB.1. Collectively, our data (**Figure 4**) demonstrate that time-evolving contributions for mutation sites identified by ICA-Var were consistent with the clinical emergence of Delta, Omicron, and XBB.1 variants. These results further solidify the foundation for the proposed pipeline, indicating its potential to identify novel mutation patterns that may lead to the emergence of new variants.

### Discovery of potential novel variants

Upon cross-referencing with dominant mutation sites in 15 VoCs (18 VoCs in **Figure 1B**, excluding emerging variants EG.5, HV.1, and BA.2.86), a set of 113 mutations sites emerged as potential novel mutations. Using a hierarchical clustering algorithm with ward distance, six clusters were obtained at a cut-off ward distance of 18 (**Figure 5A, Methods**). Among these clusters, cluster 2, 3, 4, and 5 showed overlapping mutation sites with emerging variants in late 2023 (bottom table in **Figure 5A**). Using cluster 3 as an example, we observed two sets of co-varying patterns after 06/2023 (dashed orange boxes in **Figure 5B**), both were overlapping with dominant mutations of EG.5 and HV.1. Furthermore, there were no overlapping mutations between cluster 1 or 6 with known mutation sites in emerging variants in late 2023 (bottom table in **Figure 5A**). Co-varying patterns after 2023/08 were evident for mutation sites in cluster 1 (**Figure 5D**). For these eight mutations, we verified the presence of these sites in clinical sequencing data from GISAID. Our analysis revealed that these mutations had been infrequently reported in any clinical samples (**Supplementary Figure 3**). Hence, these mutations could potentially lead to the emergence of novel SARS-CoV-2 variants and warrant close monitoring, pending clinical testing.

**Figure 5.**
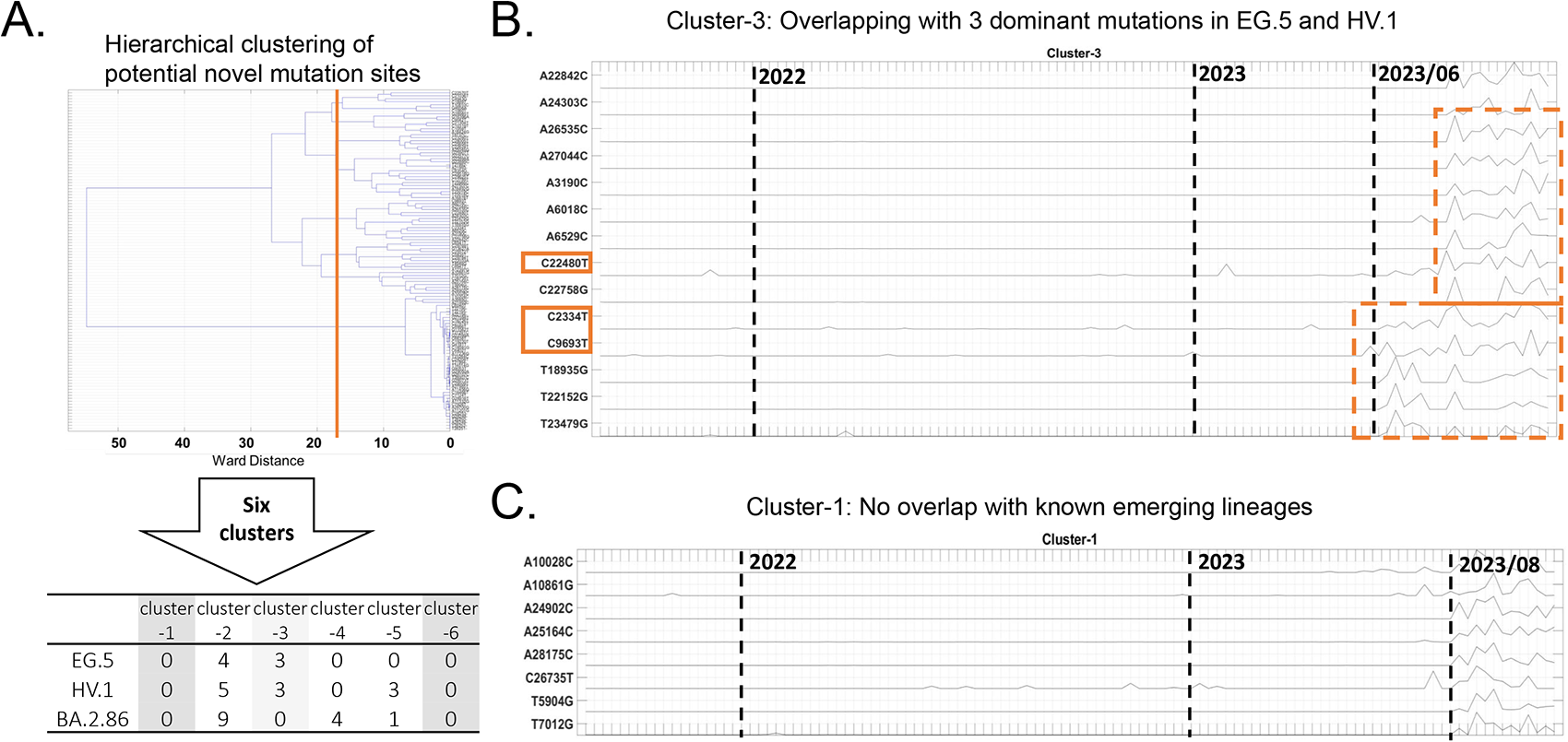
Potential novel mutation patterns. **(A)** Hierarchical clustering leads to six clusters at 113 potential novel mutation sites. Clusters 2, 3, 4, and 5 have overlapping mutation sites with emerging variants EG.5, HV.1, and BA.2.86. Cluster 1 and 6 show no overlapping mutation sites with known variants, and therefore, are more likely to give rise to novel lineages. **(B)** Co-varying patterns of mutation sites in cluster 3 show major fluctuating contributions after June 2023, consistent with EG.5 and HV.1 variants. **(C)** Co-varying patterns of mutation sites in cluster 1, with major fluctuating contributions after August 2023.

## Discussion

Wastewater-based epidemiology (WBE) offers a unique opportunity to monitor the emergence and spread of SARS-CoV-2 variants at the population level. Our proposed pipeline demonstrates early detection of the SARS-CoV-2 VoCs in wastewater preceding identification in clinical data. We further show the spatial and temporal dynamics of most emerging SARS-CoV-2 VoCs transitioning from urban to rural areas. Leveraging the data-driven nature of our proposed pipeline, ICA-Var identifies modules of mutations in the SARS-CoV-2 genome that are consistent with parallel time-changing patterns, and consequently gave rise to VoCs from August 2021 to November 2023. The proposed method offers an opportunity to identify mutation sites that are occurring simultaneously and could lead to potential novel variants, even in the absence of clinical data. Importantly, ICA-Var can also take advantage of the dual regression feature to associate an identified group source with a limited number of recently collected samples.

### Enhanced sensitivity and specificity in SARS-CoV-2 VoC detection

Wastewater samples are a composite of multiple clinical genomes spanning a local community at a given time point^40^. COVID-19 clinical testing and reports indicate that certain VoCs were dominant at specific time points from August 2021 to November 2023^41–43^. Our method, ICA-Var enables the separation of multiple genomic signals into independent sources^44^. Utilizing a significant number of wastewater samples spanning this timeframe, retrospective ICA can uncover the original mutation profile, each representing an individual or a set of similar VoCs spanning communities at different time points. Notable, ICA-Var can handle non-Gaussian and non-linearly mixed signals, operates without the need for prior knowledge, and performs blind source separation^44^. These inherent properties make ICA robust to our real-world application of de-mixing wastewater samples, enabling the identification of clinically-relevant VoCs.

Following group ICA, we performed the dual-regression analysis to re-associate the original source with weekly samples to investigate and characterize the signals from ICA within each week. Previous studies have applied the dual-regression method in functional magnetic resonance imaging data analysis to associate group networks (i.e., sources) identified by ICA with individual brain maps^45–48^. Following the same concept, we adapted the dual-regression method for our approach to project group sources back onto weekly samples. This step allows us to enhance the specificity of the group signals and provides accurate localization of mutation patterns in weekly samples. It also leads to enhanced interpretability when comparing against dominant mutations from each VoC derived from clinical sequencing data.

Collectively, as compared to the state-of-the-art Freyja tool that analyzes each individual wastewater sample with a univariate approach, the proposed pipeline boosts the statistical power and enables the earlier and more accurate detection of each VoC (**Figure 1** and **Figure 3**). The intention to incorporate deletion information in our proposed analyses also contributes to the enhanced sensitivity and accuracy (**Supplementary Figure 2**). Earlier detection of VoCs from wastewater data then enables public health authorities to implement timely and targeted interventions to mitigate the spread of the virus^40^.

### ICA-Var does not require clinical data to identify a novel variant

Wastewater monitoring and clinical sequencing data of SARS-CoV-2 can provide a comprehensive understanding of the emergence, spread and prevalence of a virus^33,49,50^. A time-dependent and accurate reference barcode for circulating VoCs are often required to identify emerging variants^34,35^. Therefore, these methods (such as Freyja and COJAC) are potentially restricted from identifying or forecasting potential novel variants in the absence of clinical sequencing data and require a “correct” barcode of circulating VoCs for accurate detection.

As a data-driven approach, our proposed pipeline (**Figure 1**) recognizes mutation sites within the SARS-CoV-2 genome through the identification of co-varying and time-evolving patterns in group sources. This crucial step allows us to identify contributing mutation sites for various VoCs in wastewater at different time points from August 2021 to November 2023, without any prior knowledge of circulating VoCs (**Figure 4**). The proposed pipeline additionally identifies co-varying mutation patterns that are more recent and contribute to emerging group sources that have not been reported in clinical sequencing data (**Figure 5**). Therefore, these mutation sites could potentially give rise to novel SARS-CoV-2 variants.

### VoCs spread from urban to rural areas

Besides methodological developments, our study shows that each SARS-CoV-2 VoC is in general first detected in wastewater samples from urban areas and later in wastewater samples from rural areas (**Figure 3**). This observation is in concordance with a previous report on COVID-19 epidemic dynamics in the United States^5^. In addition to highlighting these dynamic patterns, our results underscore the feasibility of monitoring SARS-CoV-2 VoCs through wastewater samples obtained from rural areas. Given reports that residents in rural locations are at a higher risk for disease and often lack healthcare resources^7^, WBE provides a practical and effective way to monitor the disease emergence and estimate the disease spread and prevalence.

### Limitations

As a data-driven method, ICA-Var requires a significant number of samples with high genome coverage and depth to produce stable results. Therefore, the proposed pipeline may not be suitable for scenarios with a limited number of wastewater samples or if sequencing metrics indicate genome coverage below 50% and low sequencing depth (i.e., less than 10 reads per sequenced base). Moreover, one assumption of ICA-Var is that sources are independent and linearly separable. In our application, the independence of underlying signals comes from different dominant mutation sites for various VoCs and different dominance of VoCs at different time points. Both conditions demand a relatively large number of wastewater samples to generate meaningful results. The multivariate nature of the proposed pipeline further restricts its application to detect the presence of VoCs in a single wastewater sample. Despite the re-association of group sources with individual samples during the dual-regression step, the regression nature constrains the algorithm’s stability for single-sample analyses. As a result, the proposed method can only determine the existence of VoCs within a timeframe from multiple samples, in our case, multiple samples from various wastewater sampling locations in southern Nevada.

Finally, the proposed pipeline cannot estimate the abundance of each VoC from the wastewater sample. This limitation arises from ICA-Var’s inability to distinguish between signal and noise in mixed data, and its treatment of each source with equal weight. While the former may not pose a significant concern in our application, given the bioinformatics processing pipeline’s retention of relevant SARS-CoV-2 signals, the latter impedes our ability to discern the abundance or significance of each identified source.

## Methods

### Wastewater sample collection, processing, and sequencing

A total of 3,659 wastewater samples were collected from urban and rural locations in Southern Nevada (detailed in **Supplementary Table 1**) from August 2021 to November 2023. After collection, samples were placed on ice in the field and stored under refrigeration until processing (hold time < 36 h). Nucleic acids from wastewater samples were isolated using the Promega Wizard Enviro Total Nucleic Acid Kit (Cat #A2991) following the manufacturer’s protocol. In addition, we modified the Promega protocol by lysing wastewater with the protease solution and binding free nucleic acids using NucleoMag Beads from Macherey-Nagel (Cat #744970). Total RNA (>10ng) was processed for first-strand cDNA synthesis using the LunaScript RT SuperMix Kit (New England BioLabs). Amplicon-based sequencing libraries were constructed using the CleanPlex SARS-CoV-2 FLEX Panel from Paragon Genomics. Libraries were sequenced on an Illumina NextSeq 500 or NextSeq 1000 platform with 300 cycle flow cells.

### Wastewater sequence data processing

Processing of sequencing data followed a modification of our previously published pipeline^18^. Briefly, upon sequencing, Illumina adapter sequences were trimmed from read pairs using cutadapt version 4.2^51^. Sequencing reads were then mapped to the SARS-CoV-2 reference genome (NC_045512.2) using bwa mem, version 0.7.17-r1188^52^. Paragon Genomics CleanPlex SARS-CoV-2 FLEX tiled-amplicon primers were trimmed from the aligned reads using fgbio TrimPrimers version 2.1.0 in hard-clip mode. Variants were called by iVar variants v1.4.1^39^ using mutation sites with alternative allele frequencies with respect to the reference Wuhan SARS-CoV-2 genome^53^, Genome coverage and read depth were calculated using samtools v1.16.1^54^. Strict quality control (QC) was enforced as only wastewater samples with 50x depth covering more than 80% of SARS-CoV-2 genome were retained in the following analyses. Collectively, a total of 1,385 samples, from August 2021 to November 2023, covering 59,422 mutation sites of SARS-Cov-2 variants were used for the following analyses (**Supplementary** Figure 1C-D).

### Public health sample analyses

Public health samples were processed and sequenced at the Southern Nevada Public Health Laboratory (SNPHL) as part of the Southern Nevada Health District’s surveillance of the COVID-19 pandemic. Visual inspection of sequencing reads using the Integrative Genomics Viewer (IGV) was performed to assess whether mutations had sufficient sequencing support. A TheiaCoV_Illumina_PE workflow using Nextclade version 2.14.0 was used to assign lineages.

### Retrospective independent component analysis of Variants (ICA-Var)

Mathematically, let *Y* ∈ ℝ^1,385×59,422^ denote the 59,422 mutation frequencies (i.e., the proportion of reads at a site that contains the mutation) from 1,385 wastewater samples. Since wastewater samples are aggregations of genomes from multiple infected individuals with various virus lineages, *Y* could be considered as a multivariate mixed signal of SARS-CoV-2 variants spanning the local community. The data-driven ICA approach separates this multivariate signal into additive subcomponents^44^: *Y* = *AS*, where *A* ∈ ℝ^1,385×*n_ica_*^ denotes the mixing matrix and *S* ∈ ℝ^n_ica_×59,422^ represents the source matrix (**Figure 1A**, shaded grey box). In ICA-Var, the number of ICA components (*n_ica_*) was determined from the minimum description length criterion, and fastICA algorithm was utilized to perform ICA. In our analysis, ICA was repeated 50 times with different initial values and components from each run were clustered and visualized^55^. Only reliable estimates corresponding to tight clusters were retained as final sources (*S*, **Supplementary Figure 1A**).

The original ICA method assumes that all sources (i.e., subcomponents ***S***) are non-Gaussian and that the sources are statistically independent from one another^44^. In analyzing our wastewater samples using ICA-Var, the independence comes mostly from different mutation patterns and various circulating windows of time for each VoCs^41–43^. The sparsity in *Y* contributes to the non-Gaussianness. In this case, the source matrix could represent a co-varying mutation pattern in different time windows, and could therefore serve as data-driven reference barcodes for mutation frequencies co-existing in wastewater samples. From this perspective, the ICA-Var pipeline can be considered as running a multivariate regression and determining the design matrix (***S,*** i.e., reference barcodes) from the data under the constraint of independence of the sources. In our study, we conducted a retrospective analysis using ICA-Var on 1,385 wastewater samples spanning from August 2021 to November 2023. We predicted that the ICA sources could capture the evolving dominant SARS-CoV-2 VoCs over time, each characterized by unique determinant mutation patterns.

Next, we identified a set of contributing mutations (i.e., significant mutations) for each source in *S*. We selected mutation sites with values exceeding the mean ± 2 standard deviations (i.e., 4.55% of all mutation sites) in each row of ***S*** as contributing mutations^48^. A binary matrix ^*Ŝ*^ was then computed to retain only these contributing mutations (**Supplementary Figure 1B**). The pipeline developed in this manuscript and the data used to generate the results are available at https://github.com/zhuangx15/ICAvar.

### Dual-regression to back-project source matrix onto weekly wastewater samples

To further determine the dominant mutations and VoCs for each week, we performed a dual-regression analysis^45^ to project the ICA source matrix (***S***) back onto weekly wastewater samples (**Figure 1A**, shaded grey boxes). The term “dual-regression” stems from the utilization of two regression procedures employed to estimate source and de-mixing dynamics for each week against the original data. More specifically, let *Y_i_* ∈ ℝ^*Nsample_i_*×59,422^ denote mutation frequencies for *Nsample_i_* samples in the *i^th^* week, we then, 1) used the all-sample source matrix (***S***) as a set of source regressors in a general linear model (GLM), to find week-specific de-mixing dynamics (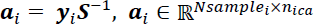) associated with all-sample source matrix (***S***); and 2) used week-specific de-mixing dynamics (*a_i_*) as a set of regressors in a second GLM, to find the week-specific source matrix (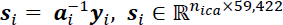) that were still associated with the all-sample source matrix (***S***). This process yields pairs of estimates forming a dual space, jointly providing the best approximation for the original all-sample ICA source matrix in each weekly sample. In summary, we obtained dual-regressed week-specific source matrix *ŝ*_*i*_ for 113 weeks from August 2021 to November 2023.

### ICA source matrix annotation to SARs-CoV-2 VoCs

To delineate the VoCs each week, we annotated our dual-regressed ICA source matrix *s_i_* by comparing them against the known mutations in VoCs from clinical SARS-CoV-2 sequencing data (**Figure 1A**, bottom row). Next, we focused on 18 VoCs that have either been or were circulating in Southern Nevada between 2021 and 2023. Due to the potential shared dominant mutations among VoCs during evolution, a hierarchical structure was formed from these 18 VoCs based on the phylogenetic tree from www.covspectrum.org (**Figure 1B**, top panel). Dominant mutation sites for each VoC were determined as follows: 1) mutations with more than 90% prevalence among clinical sequences reported at www.covspectrum.org were retained; 2) for lineages in level 2, 3, and 4 of the hierarchical tree, mutations that existed in their higher level VoCs were excluded to maintain a unique determinant mutation set (**Figure 1A**, button row); and 3) mutations with both substitutions and deletions were included in step 1) and 2). The number of dominant mutations of each VoC were listed in parentheses in **Figure 1B**, top panel.

We next binarized each row of week-specific source matrix (*s_i_*) by keeping only mutations with values greater than mean ± 2 standard deviations, as these mutations were contributing significantly towards the source (*ŝ*_*i*_). We annotated the binarized week-specific source matrix (*ŝ*_*i*_) using the dominant mutations from the VoCs by computing the following six matrices: 1) Spearman’s rank correlation coefficient (ρ); 2) sensitivity; 3) specificity; 4) area under the receiver operating characteristic curve (AUC); 5) F1 score; and 6) Jaccard Index (JI). As shown in the dendrogram of these six matrices (**Supplementary Figure 4C**), JI and F1score, AUC and sensitivity were highly similar to each other, respectively. The measure of specificity is dependent on the count of non-dominant mutations within each VoC. This count is arbitrarily determined and influences the number of dominant mutations observed in other VoCs. Therefore, we established our annotation criteria using the F1 score, sensitivity, and the Spearman’s correlation values (**Supplementary Figure 4B**), and further based on hierarchical levels and number of determinant mutations of each VoC (detailed in **Figure 1B**).

We compared VoC annotations of the proposed pipeline against results from the state-of-the-art tool Freyja^34^ (version 1.4.5, **Figure 1A**, dashed boxes). For this comparison, Freyja was retrospectively and independently applied to each of the 1,385 samples, utilizing a barcode comprising 18 VoCs, generated in October 2023. We organized samples into individual weeks, ranging from August 2021 to November 2023. In each week, if the results from Freyja indicated that any wastewater sample contained a VoC with an abundance exceeding 15%, we considered this VoC as detected by Freyja in that specific week **(Supplement Figure 4A)**.

### Potential novel mutations

Given that the identification of dominant mutation sites did not necessitate prior knowledge of reference barcodes for VoCs, ICA-Var provides a distinctive approach to discern emerging mutations. This capability extends to contributions that might give rise to novel lineages across local communities, even in the absence of clinical sequencing data. From this perspective, we focused on capturing the time-evolving contributions of significant mutations in each week-specific source matrix (**Figure 1A**, top row), using: 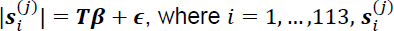, denoted the source values for *j^th^* contributing mutations in *i^th^* week, ***T*** denoted a time vector for 113 weeks from August 2021 to November 2023, and β represented the time-evolving effect of each contributing mutation. Since flipping signs of the de-mixing matrix in ICA would result in a flipping sign of the source matrix^44^, we focused on the amplitude (absolute value) of each week-specific source (*s_i_*).

A significant β indicated a critical time-changing contribution for this mutation from 2021 to 2023. As a proof of concept, we cross-checked mutations with significant β against known mutations in Delta (B.1.617.2), Omicron (BA.1), and more recent XBB.1 variants, and examined their time-evolving contributions. Following clinical reports, mutations in Delta variant (B.1.617.2) should demonstrate significant contributions in 2021, mutations in Omicron variant (BA.1) should demonstrate significant contributions from late 2021 to 2022, and mutations in XBB.1 variants should demonstrate significant contributions after late 2022.

Subsequently, we refined our pool of potential novel mutations by cross-referencing known SARS-CoV-2 variants with mutations exhibiting significant time-evolving contributions. Among the mutations retained, those demonstrating emerging contributions in more recent weeks were identified as candidates with the potential to give rise to novel lineages. To delve deeper, our focus extended to their time-evolving contributions over recent three months, from August 2023 to October 2023. We further performed a hierarchical clustering on these mutations with contributions in the three months as features, to identify co-varying patterns among these mutations. To validate the identified co-varying patterns, we examined co-varying patterns of mutations within each cluster, and cross-referenced them with dominant mutations from the emerging VoCs EG.5, HV.1, and BA.2.

## Data Availability

All data produced in the present study are available upon reasonable request to the authors

---

**Supplementary Figure 1.** Summary statistics of samples. **(A)** Independent component analysis (ICA) source matrix (*S*). **(B)** Most contributing mutations (top 5%) in each ICA source (^*S*^^). These mutation sites were focused on evaluating time-evolving contributions. **(C).** Wastewater sample coverage at 50x depth from August 2021 to November 2023. Each line represents a sequencing run and color indicates SARS-CoV-2 genome coverage. **(D)** Number of weekly wastewater samples passing quality control (50x depth covers >80% of SARs-CoV-2 genome) from August 2021 till November 2023. Urban and rural wastewater samples are plotted separately.

**Supplementary Figure 2.** Early (or simultaneous) detection of VoCs (**A**) BA.2, BQ.1, (**B**) BA.4, (**C**) BA.5, (**D**) BF.7, (**E**) XBB.1, XBB.1.5, (**F**) XBB.1.9, XBB.2.3, and (**G**) FL.1.5.1 in the proposed independent component analysis pipeline (ICA). The x-axis represents each determinant mutation for the variant and the y-axis represents each individual sample at the first detection date. The colors represent the alternative allele frequencies. Looking at the y-axis (for each sample), if Freyja outputs an abundance, the abundance value will be listed after the sample name. An asterisk (*) before the sample name indicates that the variant could be detected in the sample following the proposed criteria. For each VoC, dominant mutation sites represented by deletions are highlighted in orange boxes.

**Supplementary Figure 3.** Manual cross-refencing of mutation sites in cluster 1 for Figure 5 with clinical sequencing reports in GISAID. According to clinical reports, all eight mutations are currently not dominant in any variant.

**Supplementary Figure 4.** A comparison of detection matrices between Freyja and ICA-Var. **(A)** Maximum Freyja abundance for samples in each week. Final detection criteria in Figure 1C were set as maximum Freyja abundance greater than 15% (0.15). **(B)** Detection matrices by the proposed ICA method based on three different criteria: Spearman’s correlation value (top panel), sensitivity (middle panel) and F1score (last panel). **(C)** Hierarchical clustering results for six detection criteria computed from the proposed methods. Jaccard index and F1 score are highly similar, sensitivity and area under the ROC curve are highly correlated, and Spearman’s correlation is a unique criterion. Therefore, we utilized Spearman’s correlation, sensitivity, and F1score to establish our detection criteria in the ICA-Var pipeline (Figure 1B).

## Acknowledgments

VV, ECO are supported by NIH grants: GM103440 and MH109706 and a CARES Act grant from the Nevada Governor’s Office of Economic Development. VV, CL, DG, HK, and ECO are supported by Grant Number NH75OT000057-01-00 from the Centers for Disease Control and Prevention. The project contents are solely the responsibility of the authors and do not necessarily represent the official views of the Centers for Disease Control and Prevention. DPM was supported by the Nevada Water Resources Research Institute/USGS under Grant/Cooperative Agreement No. G21AP10578 through the Division of Hydrologic Sciences at Desert Research Institute. We would like to acknowledge personnel at DRI and the collaborating wastewater agencies for their assistance with sample logistics and data access. Special thanks to Daniel Fischer, Latoya Blanche, LeAnna Risso and Alycia S. Ybarra; to Sean Twomey, James Eason, Bill Coates, Brian Magna, Jose Rodriguez, George Veliz, Deborah Woodland; and to Chad Marchand, Teresa Gomez, and Jeremy Singleton for contributing samples from Wastewater Treatment Plants. We additionally thank Dr. Mira Han for her time reviewing this manuscript.

